# SEED: Symptom Extraction from English Social Media Posts using Deep Learning and Transfer Learning

**DOI:** 10.1101/2021.02.09.21251454

**Authors:** Arjun Magge, Davy Weissenbacher, Karen O’Connor, Matthew Scotch, Graciela Gonzalez-Hernandez

## Abstract

The increase of social media usage across the globe has fueled efforts in digital epidemiology for mining valuable information such as medication use, adverse drug effects and reports of viral infections that directly and indirectly affect population health. Such specific information can, however, be scarce, hard to find, and mostly expressed in very colloquial language. In this work, we focus on a fundamental problem that enables social media mining for disease monitoring. We present and make available SEED, a natural language processing approach to detect symptom and disease mentions from social media data obtained from platforms such as Twitter and DailyStrength and to normalize them into UMLS terminology. Using multi-corpus training and deep learning models, the tool achieves an overall F1 score of 0.86 and 0.72 on DailyStrength and balanced Twitter datasets, significantly improving over previous approaches on the same datasets. We apply the tool on Twitter posts that report COVID19 symptoms, particularly to quantify whether the SEED system can extract symptoms absent in the training data. The study results also draw attention to the potential of multi-corpus training for performance improvements and the need for continuous training on newly obtained data for consistent performance amidst the ever-changing nature of the social media vocabulary.

## 1 Introduction

Social media mining has been suggested as a promising approach in digital epidemiology to supplement traditional methods of monitoring public health. Social media posts can reveal valuable information about early symptoms from emergent infectious diseases [1, 2], medication use, abuse [3], and adherence [4], as well as adverse drug effects/events/reactions (ADEs/ADRs) [5][6] and environmental exposures that may have a significant impact on human health. While the noisy nature of the conversations (due to misspellings, informal syntax and colloquial language, for example) offers technical challenges for natural language processing methods, additional hurdles for data analysis also exist, such as selection bias in social media population and the relative scarcity of relevant postings. If not addressed, these biases could impede deriving signals for health policy and other public health interventions. Despite these challenges, through systematic methods for processing and analysis, social media has been shown to be a promising resource complementary to established data sources [7, 8, 9]. Our prior work has addressed some of these challenges, notably, for detecting adverse drug events reported on Twitter [6], for finding specific cohorts on Twitter for observational studies [10, 11], and for finding reports of switching or stopping medications [4]. Here, we extend our prior methods to focus on the identification of mentions of disease symptoms and their mapping to a standardized vocabulary. This represents an important building block for health monitoring efforts on medication safety, disease progression, or infectious disease spread.

A vast majority of prior work in this realm has focused on detecting symptoms around some other end-goal, particularly on detecting ADEs/ADRs [12, 5, 13, 6] from Twitter, Facebook, Instagram, and other forums [14, 15, 16]. More recently, social media text classifiers were developed for monitoring symptoms related to COVID-19 [17, 18, 19]. There are benefits to narrowing a data mining approach around a particular disease, as it bounds the annotation effort and can have immediate potential applications. However, the narrow approach fails to address the task of finding any and all symptoms mentioned by users, which is of particular relevance for epidemiological studies that use postings by a user or set of users over time. To the best of our knowledge, generalized extraction from social media of any and all symptoms mentioned has not been attempted. Furthermore, no effort has deployed a full pipeline of the needed mining tasks (text classification, named entity recognition, and symptom normalization) as we propose. Figure 1 shows examples of mentions of symptoms in social media posts processed in this manner.

**Figure 1:**
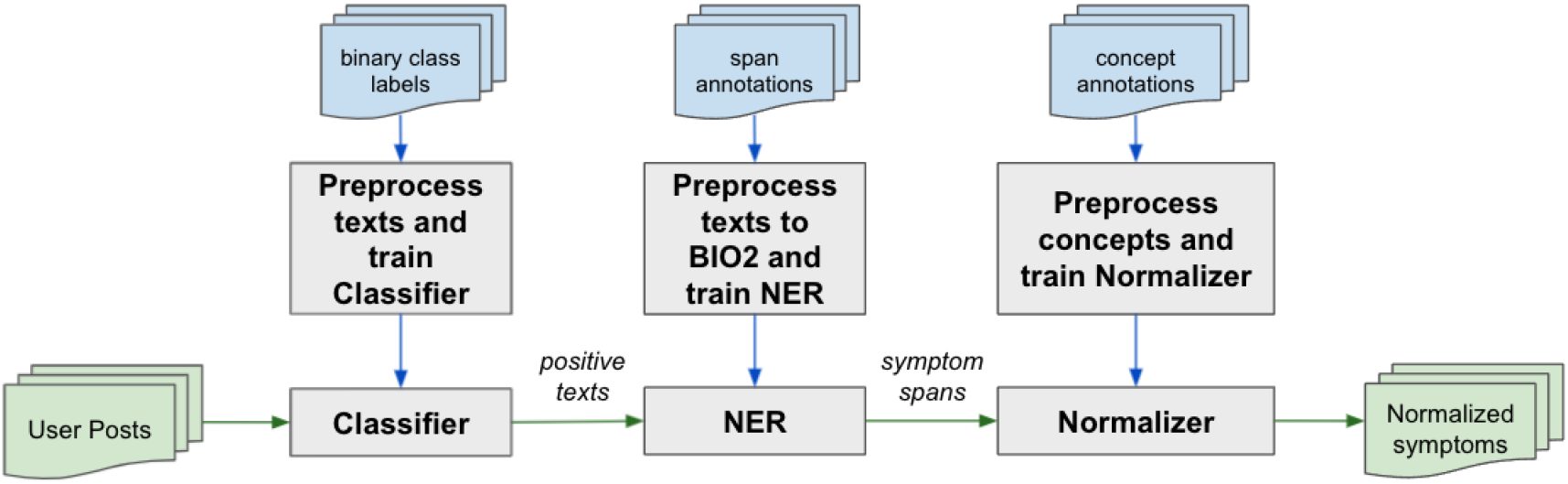
Training and inference components of the SEED tool used for extracting and normalizing symptom mentions in social media posts.

It is important to note that, in this work, we are specifically focusing and tuning our proposed approach to the detection, extraction, and normalization of mentions of *symptoms* rather than both *symptoms* and named *diseases*. The two classes are, by nature, ambiguous, and even the definition of what constitutes a ‘disease’ (or illness or syndrome) has proven controversial [20]. Addressing the task for disease presents at this point additional challenges, as the availability of annotated corpora is very limited for that class, and is poorly differentiated. In general, a symptom is defined as abnormalities noted by the patient that are indicative of a potential disease or condition [21]. Nonetheless, in this work, we do include signs, which are symptoms that can be recorded in a clinical setting. For a distinction of the classes, we refer to the Human Disease Ontology [22] (which has a parent class for disease, and one for symptom), the National Cancer Institute Thesaurus [23] (which includes a single parent class for signs and symptoms), and Ontology for General Medical Science (OGMS).

We present the first version of a social media mining tool (SEED) to identify, extract, and normalize to UMLS concept identifiers mentions of symptoms in tweets. We demonstrate the generalizability of the extraction tool and achieve state-of-the-art performance for symptom extraction using multi-corpus training. Importantly, SEED includes the first publicly available social media normalization system for symptom mention mapping to the UMLS. The SEED toolkit is publicly available at https://healthlanguageprocessing.org/pubs/seed.

## 2 Related Work

Various methods and datasets have been developed in the past decade for the extraction of medical concepts from social media. We outline the most salient ones here, noting their relevance to our work.

The *CADEC* corpus [24] is a comprehensive dataset created to facilitate the development of adverse event detection in social media. The *CADEC* dataset includes span, relation and medical concept annotations for texts collected from AskAPatient. Although the concepts annotated include *symptoms* and *diseases*, their definition of both is simply *a reason to take the drug*. Thus, CADEC uses a narrower definition of symptom (and disease) in the context of a drug, rather than in its general sense as we aim to do here. Many automatic systems have been developed using this corpora [25, 26], but almost all of them have either focused on ADE mining or concept normalization [27]. Other datasets such as *Micromed* [28] and *TwiMed* [29] that were based on tweets include annotations for symptoms, diseases and pharmaceutical products. In these works, symptoms were annotated in the tweets even when they may not have been experienced by the user. *Micromed* and *TwiMed* were small annotated datasets when they were first created, and the number of available tweets have since significantly diminished due to user and tweet deletions. The *Micromed* corpus originally contained around 1300 annotated tweets, but only 665 were available at the time of publication of the present work. Similarly, only 546 from the original 1000 from the *TwiMed* corpus were available.

Many studies have looked at extracting symptoms from social media posts as they relate to a certain disease. Focusing on depression related symptoms only, Yadivar et al [30] explored using a seeded Latent Dirichlet Allocation (LDA) topic categories, in order to address the limitations of lexical matching approaches, with terms related to depression creating a semi supervised topic modeling over time (ssToT) probabilistic model. They achieved an accuracy of 68% detecting depression related symptoms in tweets. Expanding on prior work [31] that extracted symptoms based on its frequency and cosine similarity to the term “depression”, Ma et. al., [32] developed techniques to build a semantic graph that displays the relationship between the specific term and its entities. The limitation to both these approaches is they are dependent on terminology related to a specific disease and are not readily generalizable.

More recent studies presented methods to extract COVID-19 symptoms from Twitter. Luo et al, [33] integrated syntactic dependencies and semantic relations in their deep language model developed to extract symptoms from the free text of clinical notes. Their model outperformed performed baseline models including MetaMap, BiLSTM+CRF and BERT models on clinical notes. To test the transferability of the model, they ran the model on a Twitter dataset that contained mentions of COVID-19. The performance against the Twitter evaluation set achieved an F1-score of 0.86. However the evaluation set was very small, containing only 200 tweets of which only 49 contained symptoms. Their error analysis highlighted that the model may not extract expressions of symptoms not typically found in clinical notes. Also in the COVID-19 context, Hernandez et al, [34] evaluated several annotation frameworks: SciSpaCy, MedaCy, MedSpaCy and the Social Media Miner Toolkit (SMMT) tagger, to automatically extract biomedical entities from COVID-19 related tweets without the need for manual annotation for training. They evaluated against the manually annotated dataset created in [35] finding the SMMT tagger had the highest overlap of automatic symptom/condition detection with the gold standard with 71.9% overlap. Lian et al, [36] identified tweets containing a personal COVID-19 vaccine adverse effect experienced using a Random forest algorithm for tweet classification. A CRF algorithm was used for NER to extract the vaccine names, dose, and adverse event, achieving an F1 score of 0.77 for adverse event detection.

Non-disease or event-specific identification and extraction of symptom mentions in Twitter have tended to be performed in tandem with the identification of other medical entity mentions in the tweet, such a disease and pharmacological substances. Jimino-Yepes et al, [28] proposed Micromed, a system that uses CRF as the underlying algorithm to identify disease, symptom and pharmacological substance mention in tweets. *Micromed* combined with MetaMap achieved the best performance over baseline systems traditionally used in clinical notes, with an F1 scores for symptom extraction of 0.6578 on exact matching and 0.6788 on partial matching. Improving on this prior study, they revised their approach to use a recurrent neural network using the same data set [37]. The method improved exact match for symptoms (F1: 0.676) and a statistically significant improvement in partial matching of symptom mention (F1: 0.72). Li et al [38] proposed a adversarial transfer learning for classifying tweets with ADRs using multi-corpus training to take advantage of corpus-specific and shared features.

Batbaatar and Ryu [39] proposed a Health-Related named Entity Recognition (HNER) system based on a recurrent neural network for the extraction of disease, symptom and pharmaceutical substance mentions in Twitter. The data was collected using the keyword, “healthcare”, and was automatically annotated using QuickUMLS to extract biomedical terms from text. The BiLSTM-CRF model (using word + char + POS features) had the best performance for symptom extraction (F1:87.52). Scepanoviv et. al., [40] extracted disease, symptoms and drug names using deep learning based framework and contextual embeddings. They used benchmark data sets, CADEC (Askapatient) and Micromed (Twitter), and created a validation set using Reddit. They used a BiLSTM-CRF architecture testing several contextual embeddings to find best performance. For Twitter, the combination of contextual and word embeddings (RoBERTa and GloVe) had the best performance across all entities with an F1 score of 0.72 and an F1 score of 0.74 on symptoms.

The work we present here extends various social media mining pipelines [39, 40] including ours [6] by going beyond the identification of adverse events and creating a generic system for classifying posts containing symptoms, extracting their spans and normalizing them to UMLS CUIs. We describe in detail the training and evaluation datasets, the pipeline components and their training in the Methods section. We report the results of experiments in the Results section and discuss their implications along with system limitations and future directions in the Discussion section.

## 3 Materials and Methods

### 3.1 Datasets

In this work, we reuse openly available annotated datasets for indications and ADEs. We use a total of eight annotated datasets and resources from two variants of social media posts: (a) tweets from the social media site Twitter (Tw-NER-v1 [41], SMM4H-ADE-2021[13], Micromed[28], CADEC-Tw[24], TwiMed[29]) and (b) sentence excerpts from medical/support forums such as DailyStrength (DS-NER) [41] and AskAPatient (CADEC-AAP)[24]. These datasets contain span annotations in various categories i.e. disease, symptom, ADE and indications. Only a subset of the symptoms in the aforementioned Twitter datasets are annotated along with the drug spans when they are co-located in the posts, while other datasets do not contain drug spans.

In addition to the above datasets, we also use the MedNorm resource [42] that contains normalization identifiers for commonly found symptom expressions on social media. We use SIDER database [43] to reduce the normalization label space to include only symptoms and indications recorded in the database. We refer the readers to the original papers for details regarding data collection and annotation guidelines, and present a summary of the datasets used for experiments in this work in Table 2.

**Table 1:** Examples illustrating mentions of symptoms across categories of *Indication* (in cyan) and *ADE* (in pink) in social media posts and assignment of normalization identifiers.

| Text | Extracted Spans | Normalized Spans |
| --- | --- | --- |
| This drug didn't end up helping at all. I tried it for my mood and anxiety but gave me vertigo . :( | mood<br>anxiety<br>vertigo | 10024919: Low mood<br>10002855: Anxiety<br>10047340: Vertigo |
| Is anybody else have terrible acne problems ? | acne problems | 10000496: Acne |
| I can't see the point in taking it. It's screwing up my sleep and making me depressed . | screwing up my sleep<br>depressed | 10040995: Sleep disturbance<br>10012378: Depression |
| It kinda did ok in keeping my anxiety under control. But now I need to get my heart rate down . Oh god. | anxiety<br>need to get my heart rate down | 10002855: Anxiety<br>10019303: Heart rate increased |
| Its dangerous. It is supposed to treat depression and suicidal thoughts . I took it and got very depressed . | depression<br>suicidal thoughts<br>depressed | 10012378: Depression<br>10042458: Suicidal ideation<br>10012378: Depression |
| I had a mild case of #COVID19 during the time of the protests. I was exhausted for a few weeks, chest tightness , sore throat , run down . | exhausted<br>chest tightness<br>sore throat<br>run down | 10016256: Fatigue<br>10008469: Chest discomfort<br>10068319: Oropharyngeal pain<br>10003549: Asthenia |

**Table 2:** Summary of the datasets used for the experiments presented. We merge the NER datasets to detect symptoms by combining *ADE* and *Indication* spans. Positive column indicates the percentage of tweets containing *ADE, Indication, Symptom* or *Disease* spans among all tweets in the dataset.

| Corpus | Source | Annotations used | Training | Test | % Positive |
| --- | --- | --- | --- | --- | --- |
| **DS-NER** | DailyStrength | NER spans (ADE, Indication) | 4720 | 1559 | 32% |
| **Tw-NER** | Twitter | NER spans (ADE, Indication) | 1340 | 443 | 56% |
| **SMM4H-2020** | Twitter | MedDRA (ADE) | 1786 | 1123 | 100% |
| **HLP-ADE-v1** | Twitter | MedDRA (ADE) | 2276 | 1559 | 100% |
| **CADEC** | AskAPatient | NER spans and MedDRA (ADE, Symptom, Drug, Disease) | 1000 | 250 | 100% |
| **Micromed** | Twitter | NER spans (Symptom, Drug, Disease) | 500 | 165 | 44% |
| **Twimed** | Twitter | NER spans and UMLS (Symptom, Drug, Disease) | 400 | 145 | 82% |
| **MedNorm** | Twitter, AskAPatient | MedDRA (Symptom, Drug, Disease) | 27,979 | - | 100% |

For the classification, NER and normalization tasks, we used the off-the-shelf deep learning-based Flair framework [44] to construct and train the models. We used the component architecture from our contemporary work on extracting and normalizing ADEs mentioned in social media [6]. Prior to training, 5% of the posts in the training set are randomly selected and separated into a development set for hyperparameter tuning and choosing the best model. The training was performed on a MacBook Pro 2019 with 8 cores and 16 gigabytes of RAM.

### 3.2 Classifier

The classifier acts as a filter with the purpose of removing tweets/posts that do not contain symptoms or diseases. The classifier model was built using multi-corpus training using annotations from *DS-NER, Tw-NER* and *Micromed* datasets.

The model uses the language representation layer followed by a fully connected layer with softmax layer at its output layer. All layers including the language representation layer are fine-tuned during the training procedure. The optimal settings for the Classifier were found to be a learning rate of 0.0001 with a stochastic gradient descent optimizer. The model was trained for 15 epochs and the model with the best performance on the development set was saved.

### 3.3 Named Entity Recognizer (NER)

The objective of the NER is to identify word spans that express symptoms in text and to categorize them into their relevant type of annotation. Similar to the classifier, the NER model was built using multi-corpus training from the *DS-NER, Tw-NER, CADEC, Micromed* and *TwiMed* datasets. Each text input in the annotation dataset was tokenized using segtok and the texts containing span annotations were encoded into IOB2 format. The NER model uses the language representation layer followed by a bidirectional recurrent neural network (RNN) layer (dimension size 256) composed from gated recurrent units (GRU) which are then concatenated and passed through a fully connected layer. Outputs from the fully connected layer are then passed through a conditional random field (CRF) layer. The weights in the model are trained by the minimizing the loss computed using the negative log likelihood computed at the CRF layer. Unlike the classifier, only the fully-connected layer and output layer are fine-tuned during the training procedure.

The optimal settings for the NER were found to be a learning rate of 0.1 with a stochastic gradient descent (SGD) optimizer. The model was trained for 70 epochs and the model with the best performance on the development set was saved for testing its performance on the test sets. Among the language representation techniques available, FastText embeddings with enriched subword information [45], BERT [46] and RoBERTA [47] models were found to perform the best.

### 3.4 Normalizer

The normalizer is built based on our recent work [6] which employs a classifier that is trained using semi-supervised learning. Semi-supervised learning uses a combination of supervised and unsupervised learning i.e. the model is trained on both human annotated MedDRA identifiers and spans available from medical ontologies. Training in such a semi-supervised manner allows for discovery of symptoms not available in the training set and for better generalization capabilities. In this work, we use MedDRA’s lower level terms (LLTs) as a training set with preferred terms (PTs) as target labels. Furthermore, MedDRA term identifiers were integrated with terms from other ontologies in the UMLS metathesaurus [48] using their respective concept unique identifiers (CUIs) and included in the training dataset. MedDRA contains over 23,000 preferred term identifiers which include laboratory observations, diagnoses and tests that are mostly not considered symptoms. Hence, for the purposes of training SEED, we reduce this label space include common symptoms recorded in the SIDER database [43].

We use the training sets with available normalization annotations (MedNorm, HLP-ADE-v1, SMM4H-2020, CADEC and TwiMed) to create a multi-corpus dataset of training instances where each entry is unique. We use this dataset primarily for validation and testing purposes. Using the annotated datasets for training will create a model that capable of predicting the classes in the training set. However, we want SEED to be generalizable and capable of predicting classes that are not present in the human annotated datasets. Hence, we add entries from UMLS for creating an expanded dataset for training purposes such that additional class labels that are not part of the training procedure are included in the training procedure. These entries are used to train a classifier that uses transformer embeddings such as DistilBERT, BERT and RoBERTa. During training, the semi-supervised dataset is used completely for training until maximum accuracy is achieved on the supervised dataset.

### 3.5 Validation

To validate the performance of SEED on newer data, we extracted symptoms from a dataset created on COVID19 related terms [10]. To prepare for this task, we retrained the *Normalizer* on 20 additional supervised examples containing spans of COVID mentions e.g. *covid, covid19, coronavirus* from the latest version of MedDRA. We analyzed their results by sampling and annotating 500 tweets that were predicted by SEED to contain symptoms (*COV-Symptoms*) to look for false positives. We also analyzed 500 tweets that were predicted by SEED to not contain any symptoms (*COV-NoSymptoms*) to look for false negatives. Prior to the analysis, an expert annotator annotated symptom mentions as well as MedDRA normalization identifiers for the two sets of 500 tweets.

## 4 Results

We tested the performance of SEED on the datasets listed. For the purposes of comparison of NER with previously developed systems, we include the evaluation results for the Classifier and NER in Tables 3 and 4 respectively. We observe that multi-corpus training is largely beneficial for both the Classification and NER tasks and exceed the previous state-of-the-art performances on two (*DS-NER* and *Tw-NER*) of the three datasets. We find that there was no improvement in the performance on *Micromed* dataset under multi-corpus training for the Classification task.

**Table 3:** Comparison of single and multi-corpus training on the classification datasets using the RoBERTa-Large transformer model. Multi-corpus training shows performance improvement in datasets annotated with similar guidelines and show minor loss in other datasets.

| Training Set | Single-corpus<br>$P/R/F_1$ | Multi-corpus<br>$P/R/F_1$ |
| --- | --- | --- |
| DS-NER | 0.97/0.88/0.92 | **0.93/0.92/0.93** |
| Tw-NER | 0.76/0.98/0.86 | **0.87/0.86/0.87** |
| Micromed | **0.84/0.86/0.85** | 0.83/0.82/0.83 |

**Table 4:** Comparison of single and multi-corpus training on NER datasets. SOTA performance on the *CADEC* dataset could not be determined as other works use different entity subsets as symptoms. SOTA performance on *Micromed* was reported on the full dataset, however, as of this work, only 51% of the tweets are available.

| Training Set | SOTA<br>$P/R/F_1$ | Single-corpus<br>$P/R/F_1$ | Multi-corpus<br>$P/R/F_1$ |
| --- | --- | --- | --- |
| DS-NER | 0.86/0.78/0.82 | 0.88/0.82/0.85 | **0.88/0.84/0.86** |
| Tw-NER | 0.76/0.68/0.72 | 0.78/0.70/0.74 | **0.79/0.71/0.75** |
| CADEC | - | **0.75/0.81/0.78** | 0.74/0.80/0.77 |
| Micromed | **0.79/0.66/0.72\*** | 0.65/0.62/0.64 | 0.63/0.60/0.62 |
| Twimed | - | 0.57/0.68/0.62 | **0.58/0.68/0.63** |

For the NER task, we see improvements in the *DS-NER* and *Tw-NER* datasets, that were based on similar annotation guidelines. However, we find that the performance on *CADEC* and *Micromed* datasets decrease under multi-corpus training. When trained at the sub-entity levels such as *ADE* and *Indication* that were available on the *DS-NER* and *Tw-NER* datasets, we found that training on *DS-NER* increased the *Tw-NER* model’s performance by 13 percentage points for *ADE* and 23 percentage points for *Indication* extraction. Training on *Tw-NER* dataset had a beneficial effect for *DS-NER* model only for *ADE*. This establishes a new state-of-the-art performance over the previous state-of-the-art by the ADRMine system [5] which achieved F1= 0.82 on DailyStrength and F1 = 0.72 on Twitter datasets for *ADE* extraction, compared to our new F1 = 0.86 and F1 = 0.75, respectively. After experimenting with various forms of language representation techniques, *BERT* [46], *RoBERTA* [47] and *BERTweet* [49] models were found to perform the best with the *BERTweet* model achieving the best performance. The normalization model was evaluated on the *SMM4H-2020, HLP-ADE-v1, CADEC* and *TwiMed* datasets. On average, the cross-evaluation accuracy of the Normalizer was recorded to be 0.63. Due to lack of other comparable systems that perform extraction and normalization, a direct comparison with SEED was not possible.

## 5 Discussion

Table 5 shows selected examples of some errors resulting from the SEED system. We find that the number of errors in the pipeline are evenly spread out among the three components i.e. Classifier, Extractor and Normalizer. A vast majority of the errors were due to false negatives at the NER stage of the pipeline. While the classifier threshold was optimized for better recall, there were many tweets with symptoms mentions that were not recognized by the classifier. Unique phrases such as *eighty years old feeling* that are not likely to be present in the training stage were responsible for a vast majority of the errors by the classifier and the extractor.

**Table 5:** Error analysis performed on on the multi-corpus test dataset. Bold text represent gold standard annotations while italicized text represent predicted spans of text.

| Sentence | Error | Category |
| --- | --- | --- |
| I just woke up from a **long 13 hour nap**. Thank you Fluoxetine. | False Negative | Classifier Error |
| I just spent 85 dollars on **allergy** meds. Bought 2 of everything. | False Negative | Classifier Error |
| I have been on this drug for years. I was on 75 mg 1x a day and now 2x a day. I really didn't have any *bad side effects*. | False Positive | Classifier Error |
| In older men *impotence* has a physical cause, such as disease, *injury*, or side effects of drugs | False Positive | Extractor Error |
| today was ment to be my *chill* day and my *stress* levels are once again through the roof | False Positive | Extractor Error |
| Yesterday's trivia answer is that Pasteur was an expert in treating and preventing **rabies**, an often fatal disease. | False Negative | Extractor Error<br>Possible Annotation Error |
| I gotta feeling that imma *throw up* tonight. not feeling bad but I keep **burping** and **vomit keeps coming up** everytime. | Partial False Positive |  |
| How **nervous** was she performing for music? That was like a **blackout** experience. | False Negative | Extractor Error |
| I still have the same **eighty years old feeling** and do not know that i will ever be normal again. | False Negative | Extractor Error |
| I too am **very tired** lately and *never feel rested* upon awakening. | Partial False Positive | Extractor Error |
| My *mind is more important* than any drug | False Positive | Extractor Error |

During error analysis in the validation stage, one of the primary observations we made is that newer disease terms such as *covid, covid19, covid-19 coronavirus* were detected in only 10% of the COV-Symptoms set of tweets. We find that annotating a few tweets that mention COVID19 symptoms is beneficial when a model trained on generic symptoms runs on such a dataset. We expect that this might extend to symptoms of diseases that are not represented in the training corpora. Overall, from the *COV-Symptoms* set of annotations, 1356 symptom mentions were extracted and 390 mentions were ignored by the SEED system. Table 6 shows the most common symptoms mentioned in the *COV-Symptoms* set of tweets. The frequent symptoms such as cough, fever, sore throat, headache, and chest pain extracted from SEED have been observed among patients suffering from coronavirus[50] [51]. Among the set of 500 tweets, SEED found three instances of Ageusia (loss of sense of taste) and Anosmia (loss of sense of smell) that are known rare symptoms [50].

**Table 6:** Most frequently mentioned (normalized) symptoms in the COVID19 tweet dataset listed with an example mention. All tweets have been paraphrased to preserve anonymity.

| Normalized Symptom | Perc of all symptoms | Example of symptom mention with span highlighted |
| --- | --- | --- |
| Malaise | 13.70% | I'm **sick** and googling if I have a cold or coronavirus. |
| Cough | 11.33% | For the last 4 days I've had a runny nose and a **tickling cough** at night. |
| Pyrexia | 10.15% | Someone told me u normally dont run a **high fever** with pneumonia plus I've had a headache the whole time. |
| Pneumonia | 3.93% | My 15 year old has had a fever and dry cough for a week. Today he tested positive for **pneumonia**. |
| Influenza | 3.70% | I'm not sure yet if I will test positive for coronavirus, but I do know that I had the **flu** in mid-Dec. |
| Dyspnoea | 3.26% | So today I find out if what brought me in with **bad breathing issues** and a high fever is coronavirus. |
| Pain | 2.81% | Have someone in my house who is more than likely infected with the coronavirus. They're an er doctor and they've come down with a 102 degree fever and **body aches**. |
| Oropharyngeal pain | 2.59% | Yup. I have GERD which means that I have a **sore throat** and shortness of breath most days. |
| Fatigue | 2.15% | Since a week or so into isolation: Sore throat, GI, dry cough, low fever, **fatigue**, all mild, spread between us. |
| Headache | 1.63% | SAME AF! then my anxiety starts making me think the **headache** is coronavirus, which makes my anxiety 50x WORSE. |
| Bronchitis | 1.26% | I was denied testing multiple times because I didn't have a fever AND because I have history of **bronchitis** and pneumonia so I wasn't a priority test. |
| Asthma | 1.19% | When they tested me I had the flu, viral pneumonia, and viral sepsis. Now I am **asthmatic** and have 2 inhalers. |
| Anxiety | 1.19% | Since someone told me I was exposed to covid-19, I've realized lots of covid symptoms overlap with **anxiety**. Shortness of breath, nausea, headache, dizziness. |
| Chest pain | 0.96% | Wife had fever, **chest pain**, cough. Quarantined in our bedroom for 2 weeks. Never able to get a test. A week into the quarantine, she mentioned a weird kind of athletes foot...but more painful and burning. Looked JUST like this. #covidtoes |

Analyzing the *COV-NoSymptoms* set of tweets with expert annotations, SEED failed to detect some COVID19 related symptom mentions such as *COVID-19 pneumonia, lung opacity* and *decrease in oxygen saturation* that were found by the human annotator. Overall around 190 mentions i.e. false negatives were not found in the *COV-NoSymptoms* set of tweets.

### 5.1 Limitations and Future Work

The tool presented in this work, SEED, unlike prior work, is generic enough to be used in isolation on social media posts regardless of whether the post also includes a drug mention. However, the performance of the tool has been evaluated on datasets that have higher incidence of symptoms compared to a random sample of tweets from Twitter. Hence, for such datasets where the occurrence of symptoms are low, additional noise removal and filtering strategies such as the use of regular expressions, rules and/or retraining the supervised classifiers are recommended. Another limitation of the SEED system is that the datasets used for its training included tweets collected between 2013 and 2015. To test its performance on more contemporary data, we included a newly annotated COVID-19 dataset.

## 6 Conclusion

In this work we present a system developed using multi-corpus training to extract and normalize symptoms on social media posts. On a filtered balanced dataset, it obtained state-of-the-art performance for extracting symptoms, outscoring the SOTA ADRMine system on the Twitter and DailyStrength datasets. Equipped with state-of-the-art components and multi-corpus training, SEED achieves the best performance on classifying posts that contain symptoms, extracting mentions of symptoms and normalizing the mentions to a standardized vocabulary. We have made SEED publicly available to users as a standalone tool, a web-based demonstration interface, and an application programming interface which performs symptom extraction and normalization tasks on user submitted content.

## Data Availability

Datasets and code are available from the Health Language Processing website.

https://healthlanguageprocessing.org/pubs/seed/

## Acknowledgements

A portion of this work was done when AM was a graduate student at Arizona State University supervised by GG and MS. MedDRA® the Medical Dictionary for Regulatory Activities terminology is the international medical terminology developed under the auspices of the International Council for Harmonisation of Technical Requirements for Pharmaceuticals for Human Use (ICH). MedDRA® trademark is registered by IFPMA on behalf of ICH.

